# A Machine Learning Decision Support Tool Optimizes Whole Genome Sequencing Utilization in a Neonatal Intensive Care Unit

**DOI:** 10.1101/2024.07.05.24310008

**Authors:** Edwin F. Juarez, Bennet Peterson, Erica Sanford Kobayashi, Sheldon Gilmer, Laura E. Tobin, Brandan Schultz, Jerica Lenberg, Jeanne Carroll, Shiyu Bai-Tong, Nathaly M. Sweeney, Curtis Beebe, Lawrence Stewart, Lauren Olsen, Julie Reinke, Elizabeth A. Kiernan, Rebecca Reimers, Kristen Wigby, Chris Tackaberry, Mark Yandell, Charlotte Hobbs, Matthew N. Bainbridge

## Abstract

The Mendelian Phenotype Search Engine (MPSE), a clinical decision support tool using Natural Language Processing and Machine Learning, helped neonatologists expedite decisions to whole genome sequencing (WGS) to diagnose patients in the Neonatal Intensive Care Unit. After the MPSE was introduced, utilization of WGS increased, time to ordering WGS decreased, and WGS diagnostic yield increased.

## Main Body (Results & Discussion)

Genetic disorders are a leading cause of death and disability for infants admitted to the Neonatal Intensive Care Unit (NICU)^1^. Rapidly diagnosing the underlying cause of critical illness and initiating targeted treatment are of paramount importance given the considerable morbidity and mortality associated with NICU admission^1–6^. Rapid Precision Medicine utilizing Whole Genome Sequencing (WGS) can help identify patients with genetic disease and thus facilitate care tailored to the individual^5–13^. However, due to economic considerations and clinician familiarity with WGS, deciding which patients should receive WGS in the NICU can be challenging^6,12–15^. We hypothesized that an automated clinical decision support tool utilizing machine learning to continually reassess the appropriateness of rapid WGS (rWGS) could assist neonatologists with patient prioritization for rWGS.

A single-group study was designed to compare findings before and after the implementation of a clinical decision support tool. The clinical support tool, Mendelian Phenotype Search Engine (MPSE), was designed to utilize Machine Learning (ML) to leverage the Human Phenotype Ontology (HPO) terms to calculate scores for prioritizing patients for WGS^16^. The HPO provides a hierarchical representation of the clinical abnormalities observed in human disease, and thereby facilitates computational analysis of patient phenotypes^17,18^. Natural Language Processing (NLP) tools can identify HPO terms found in Electronic Medical Record (EMR) notes that describe patient phenotypes related to Mendelian disease, allowing for analysis via machine learning (ML)^19–22^.

We developed a software pipeline to automatically extract HPO terms from unstructured physician notes embedded within the EMR of patients recently admitted to the NICU. These HPO terms were used by the MPSE to compute a prioritization score that reflects the similarity of newly admitted NICU patients to observed phenotypes of patients within the NICU who previously received WGS^16^.

We performed this study in two phases. The objective of Phase 1 (the pre-implementation phase), was to collect baseline data on the number of babies nominated for WGS, the time to nomination, and the diagnostic yield of WGS. During this phase, MPSE scores for each patient were computed daily but were not provided to the clinical team. During Phase 2 (the implementation phase), the attending neonatologists were provided with a daily report containing MPSE scores for each NICU patient on the census. This MPSE report was presented to the neonatologists as an additional piece of information to be taken into consideration when deciding which patients should receive WGS.

Three primary outcomes were measured: 1) number and proportion of babies nominated for WGS; 2) time from admission to nomination for WGS, and; 3) diagnostic yield of WGS.

In total, 118 patients were nominated for rWGS; 27 in Phase 1 (14 weeks, 1.9 nominations/week) and 91 in Phase 2 (38 weeks, 2.4 nominations/week) (Mann-Whitney-Wilcoxon two-sided test, p=0.35); in both phases 13% of the eligible patients in the NICU were deemed by the attending physician to benefit from rWGS. Of the nominated patients, 98 patients (83%) were enrolled and underwent WGS (reasons for decline listed in Supplementary Table 1); 25 from Phase 1 (1.8 enrollments per week) and 73 from Phase 2 (1.92 enrollments per week). Enrollment rates were not significantly different between Phase 1 and Phase 2 (Mann-Whitney-Wilcoxon two-sided test, p=0.63).

Of the 99 sequenced patients, 29 received a molecular diagnosis (Supplementary Table 1), with 6 diagnoses in Phase 1 (24% diagnostic yield) and 23 in Phase 2 (32% diagnostic yield) (Fisher’s Exact test, p=0.61). Each of the diagnosed patients had at least one genetic variant consistent with their phenotype classified as pathogenic or likely pathogenic according to The American College of Medical Genetics and Genomics (ACMG) guidelines^23^.

The median time from admission to nomination decreased from 48.0 hours in phase 1 to 39.1 hours in phase 2 (18.5% reduction; Mann-Whitney-Wilcoxon two-sided test, p=0.10). This is particularly noticeable at 72 hours post admission where, in phase 2, 82% of all nominations had taken place, vs only 53% in phase 1 (Cox’s proportional hazard regression, p=0.10).

**Figure 1:**
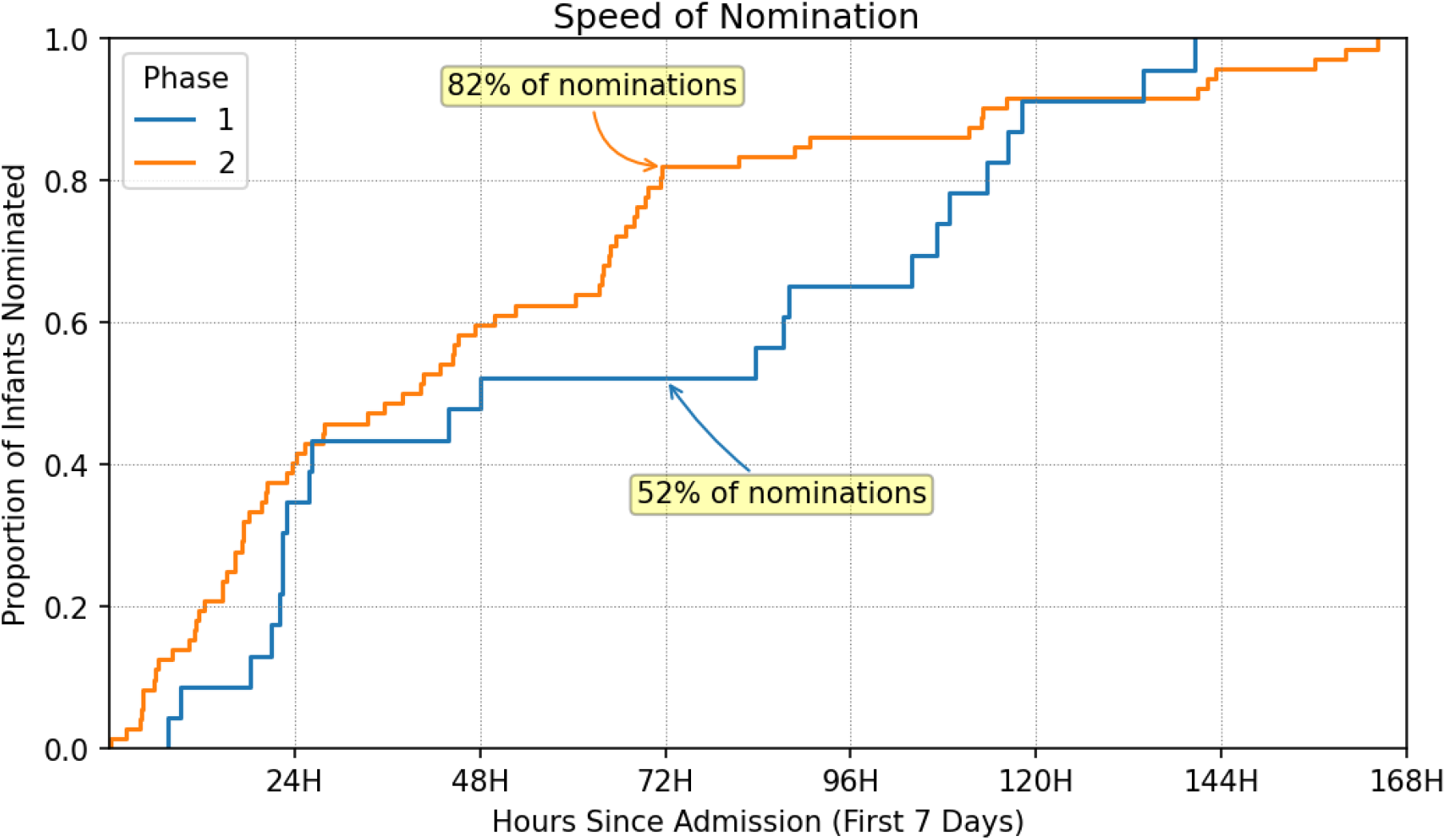
Speed of nomination curves showing the time-to-nomination for phase 1 (blue curve) and phase 2 (orange curve) patients nominated within the first 7 days of their NICU stay.

**Figure 2.**
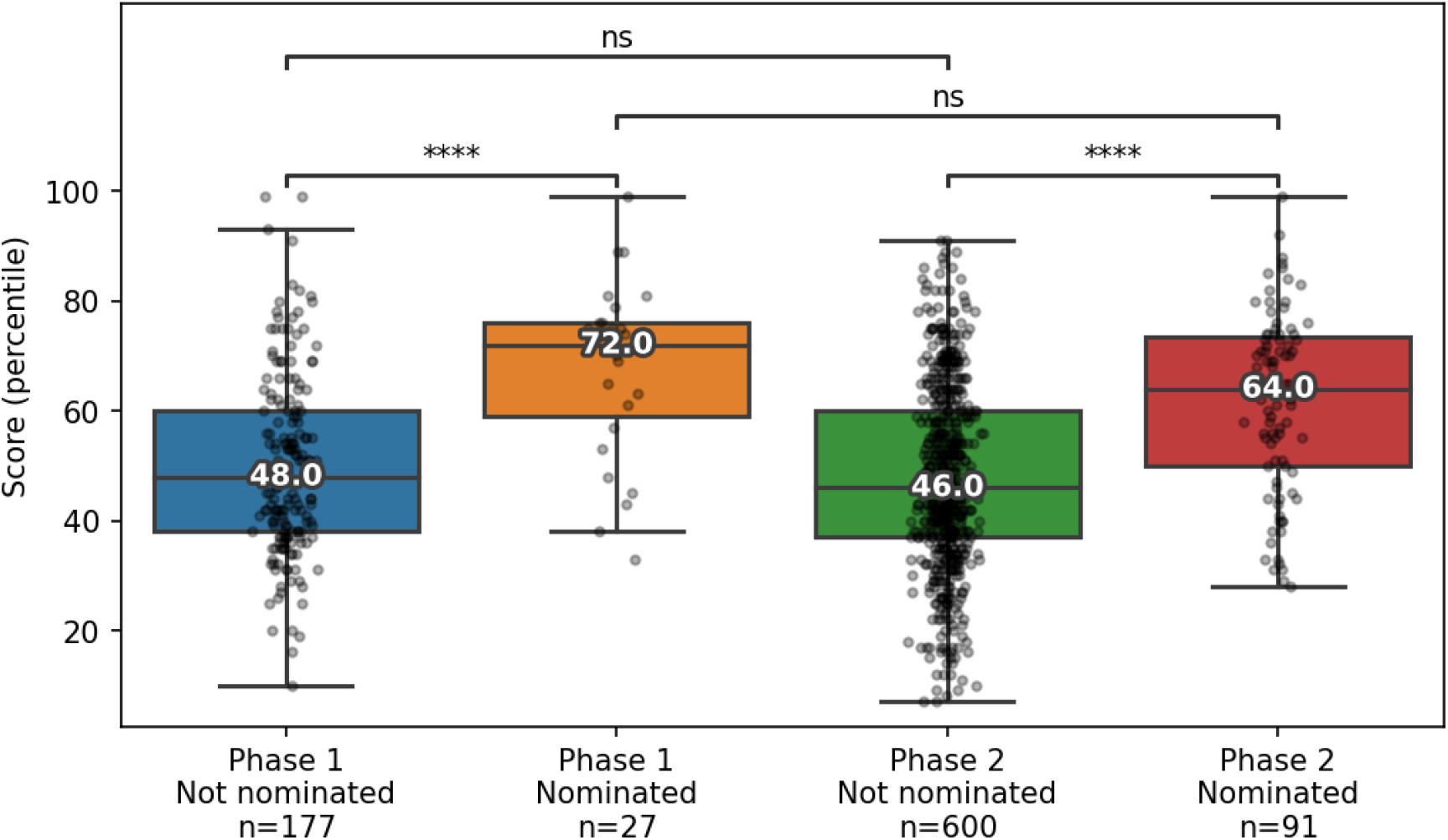
MPSE score percentiles for every patient admitted to the NICU during the duration of the study. For nominated patients, the MPSE score at the time of nomination is shown. For patients who were not nominated, the maximum MPSE score within the first seven days of their NICU admission is shown.

In both phases the MPSE scores for the nominated patients were significantly higher than the scores of the patients who were not nominated (Mann-Whitney-Wilcoxon two-sided test; Phase 1: p=1.5×10-4; Phase 2: p=4.6×10-17) and is consistent with our previous work that found that the MPSE scores of patients nominated for WGS were higher than those not nominated^16^.

Although the differences between the three primary outcomes were not statistically different between pre-implementation (Phase 1) and implementation (Phase 2) of MPSE, there was a trend towards improvement for all three primary outcomes (number and proportion of babies nominated for WGS, time from admission to nomination for WGS, and diagnostic yield of WGS) in Phase 2 when MPSE data were used to assist neonatologists’ decisions to use rWGS. Specifically, observed promising results were found regarding nomination frequency, nomination speed, and diagnostic yield after utilizing MPSE for clinical decision support. After implementing MPSE, we saw a modest but important increase in both the speed of nomination (how soon after admission does nomination occur) and weekly rate of nominations.

Importantly, increased nomination frequency and decreased elapsed time between admission and WGS nomination after introduction of MPSE were observed together with a modest increase in diagnostic yield (24% to 32%), suggesting that the increased frequency and speed of nomination did not degrade the yield of rWGS and may have improved it.

Limitations in this study include a small sample size, especially during pre-implementation and lack of long-term outcome data. These challenges should be addressed with future studies. Additionally, the established familiarity of the study site’s NICU physicians with rWGS suggests MPSE might hold greater influence in settings where Rapid Precision Medicine has not been established. Further research is needed to confirm these preliminary findings and to assess generalizability between NICUs and clinical teams.

Although statistically significant differences were not observed, likely due to limitations in sample size, these findings hold promise for future research. This study contributes to the ongoing effort to inform the design and implementation of ML tools within healthcare environments. This study demonstrates MPSE’s capability for integration into existing clinical workflows and indicates MPSE could be similarly employed at other healthcare systems.

These findings underscore the immediate impact that carefully applied clinical decision support tools harnessing NLP and machine learning can potentially have for clinicians in the intensive care unit with regards to efficiently and appropriately selecting patients for genomic sequencing.

## Methods

### Patient enrollment

This clinical prospective study was conducted in the Level IV NICU of Rady Children’s Hospital in San Diego (RCHSD). Our study was implemented in 2 phases. In each phase, attending neonatologists nominated patients for WGS following broad inclusion criteria, which included any NICU patient within the first seven days of life who was suspected of having a genetic disease or a patient with an abnormal response to therapy after the first seven days of admission to the NICU. The patient’s family provided written informed consent, and whenever possible, parent samples were also collected.

Phase 1 lasted 14 weeks (July to October 2022) and Phase 2 lasted 38 weeks (October 2022 to July 2023).

### MPSE Score Computation

The Mendelian phenotype Search Engine (MPSE) employs Human Phenotype Ontology (HPO) terms to determine the likelihood that a Mendelian condition underlies a patient’s phenotype. MPSE employs a simple, well-established approach: a Naïve Bayes (NB) classifier that has previously been published in detail by our group^16^. Briefly, MPSE uses the differences in HPO term frequencies between a collection of cases and controls to score each patient by calculating NB the log-odds ratio.

HPO-based phenotype descriptions were generated for all patients in Phases 1 and 2 by NLP analysis of clinical notes using CLiX ENRICH (Clinithink, Alpharetta, GA). A pre-trained MPSE model was then used to calculate MPSE scores for each patient^16^. In this report, MPSE score percentiles are reported to simplify interpretation.

MPSE scores and percentiles for each patient in the NICU were computed automatically every three hours during the study period. Each score’s percentile represents the position that a given score would have taken in the training cohort, thus percentiles can be compared to each other without the need to recalculate them with each new score added to the distribution.

### Statistics

Statistics were computed in Python version 3.10.2 with SciPy version 1.8.0, statannotations version 0.5.0, and lifelines version 0.27.8.

Group statistics utilize the Mann-Whitney-Wilcoxon two-sided test, the Fisher’s Exact test to compare proportions, and Cox’s proportional hazard model to compare time to nomination. For all testing, p < 0.05 was considered statistically significant.

## Data availability

De-identified data utilized in this paper is attached as supplementary material; including time from admission to nomination, MPSE score, WGS results for enrolled patients, and reasons for decline for patients who did not enroll.

## Acknowledgments

This study was funded by the Conrad Prebys Foundation, R.R. is supported by NCATS grant number K12TR004410. We are grateful to the families who participated in this study. We are also thankful to the RCHSD NICU team for their collaboration and contributions to this study.

We acknowledge the assistance that Ricky (Hung) Nguyen provided to the study by delivering the daily reports and Shauna Briscoe for serving as a project manager.

## Author contributions

C. H., M. Y., C. T., J. R. conceptualized this study design and supervised the implementation; E. J. coordinated the implementation of the software and report delivery, performed the data analyses, and wrote the first draft of the manuscript; M. B., M. Y., C. H., L. T., and C. T. revised the figures and data analyses; M. B., E. S. K., B. P., C. H., and L. T. revised earlier versions of the manuscript; E. S. K., R. R., K. W., J. C., S. B. T., and C. H. provided clinical expertise throughout the study and during manuscript revisions; M. B., C. T. and M. Y. provided supervision for data analysis and interpretations; E. J., S. G., C. B., D. D., C. T., M. Y., and C. H., designed and implemented the score report. J. L., B. S., E. K., L. O., E. S. K., R. R., K. W., and C. H. contributed to study enrollment; and J. L., B. S. returned results to patients’ families. All authors reviewed and approved the final versions of this manuscript.

